# Prevalence of Screen Addiction and its association with Screen Use Behavior & Type of Content Consumed in the general population of Mumbai & its environs

**DOI:** 10.1101/2023.02.15.23286001

**Authors:** Shirish Rao, Vidushi Gupta, Amey Ambike, Shilpa Adarkar, Pauras Mhatre, Prashant Saraf, Shreeya Raul, Raghav Paranjape, Keval Dedhia, Gayatri Inamdar, Pavan Chavan, Purva Shinde, Esha Kadam

## Abstract

**Background:** Varied prevalence of internet, gaming & smartphone addiction have been estimated across different regions. Though gaming & internet addiction have already been recognized, the emerging trend of binge-watching also demands attention. There was a need to estimate their prevalence using a uniform scale, compare addiction scores and also assess its association with content consumed and screen use behavior.

**Objective:** To find the prevalence of screen addiction and its association with screen use behaviour & type of content consumed.

**Methodology:** A cross-sectional study was conducted among 252 participants who were recruited using stratified random sampling and were interviewed using Digital Addiction Scale & a self-designed questionnaire (CVR>0.65).

**Results:** The prevalence of screen addiction was found to be 13% and found to be associated with age(p<0.01), sex(p<0.01), type of content consumed(p<0.05), device used(p<0.01), self-reported causes(p<0.01), withdrawal(p<0.01) & screen use specific psychological phenomenon(p<0.01). Addiction scores of gamers, social media users & binge watchers were comparable(p>0.05).

**Conclusion:** The prevalence of screen addiction is substantially high, particularly in the younger population. This study also highlights the strong association of screen use behavior & type of content consumed and apps used with addictive screen use. The similarity in addiction scores and associated factors also support the use of umbrella term ‘Screen Addiction’ to group all kinds of addictive screen usage.

## 1. Introduction

Over the past 30 years, a marked expansion has been observed in screen-based communication and entertainment options available to adolescents. Along with existing technologies such as television & computer, many adolescents now have easy access to, online instant messaging, social networking websites and online video streaming platforms.

As of January 2021, 4.66 billion people were active internet users across the globe – which is around 59.5% of the global population. India alone had almost 560 million online users by 2020. ^[1]^ Around 92.6% internet users access the internet through mobile devices, but the use of computers also contributes to a fair share of the internet activity, as greater than 70% of internet users residing in the larger economies of the world, go online via laptops and personal computers for quite a few of their connected activities. In the past 12 months, the global number of social media users increased by 490 million^[2]^. As of now, India has almost 365 million mobile gamers, whose numbers are expected to keep on growing rapidly^[3]^. One survey reported a huge rise of 23% in the time spent in using OTT platforms during the first national lockdown from mid-March to July. People between ages 17-35 years – which account for 49 % of India’s population – spent around 8-9 hours a day binge-watching digital content. The average time spent in accessing OTT video content on a daily basis by millennials & Gen Z in India, was 7 hours, which is nearly twice the global average (4 hours)^[4]^.

Although the last 10 years have seen a decline in traditional television viewing, the use of newer screen-based devices for watching T.V. shows and other content has risen steadily, thus leading to a net increase in average screen time. ^[5]^

Nowadays, discretionary screen time (DST), which often involves multiple devices, is the only major experience and environment that individuals are exposed to. The growing concern over this increasing amount of screen time has led to more and more physicians using the term ‘addiction’, to describe the growing number of individuals being involved in a myriad of screen activities, in a dependent and problematic manner. ^[6]^

An addiction in which any activity (like TV, gambling, gaming, and the Internet) shows signs of pathological dependence similar to that seen in any drug addiction, without actually being dependent on any particular substance, is defined as ‘behavioural addiction.’ ^[7]^.

As given by Griffiths et al. (1999) in their article titled ‘Internet Addiction: Fact or Fiction?’, there are six criteria to consider any behaviour as an addictive one i.e salience, mood modification, tolerance, withdrawal, conflict and relapse. ^[8]^

Although the term ‘screen addiction’ has been restricted to the context of video gaming, excessive messaging and social networking, this concept should essentially include the undertaking of any potentially addictive activity, which involves a screen. However, even though watching TV series and movies contributes significantly to screen exposure, it still remains neglected. ^[9]^

A meta-analysis found the global prevalence of internet addiction to be around 6%.^[10]^ In the Indian context, general population studies have shown the prevalence of internet addiction to be 1.3%.^[11]^ This increased almost 10-fold (11.8%, 8.8%, and 8%) in college-going students. ^[12, 13, 14, 15]^ While one study in India amongst health professionals reported the prevalence of severe internet addiction among dental students (2.3%), as well as other medical students (1.2%), another similar study showed the prevalence rate of internet addiction to be 9.5% among medical college students as a whole ^[16]^. The substantial variation observed in these rates may be partially accounted to the inconsistencies across the studies in the manner of quantifying internet addiction.

Research focussing on the use of screen-based media and its addictive potential has been limited in India. With the rapid advancement of screen-based options for entertainment, communication, and education, more studies are needed to assess the psychological consequences of these diverse types of content. Varied prevalence of internet, gaming & smartphone addiction have been estimated across different regions. Though gaming & internet addiction have already been recognized, the emerging trend of binge-watching demands attention. There is also scarcity of evidence with regard to associated withdrawal symptoms in binge watchers and newly emerging psychological phenomena like texting anxiety, game transfer phenomenon and para-social relationships. Hence, there was a need to estimate the prevalence of screen addiction using a uniform scale, compare addiction scores and assess its association with content consumed and screen use behavior.

## 2. Objectives

1. To find the prevalence of screen addiction and its association with screen use behaviour & type of content consumed.
2. To compare the addiction scores with respect to types of devices used & type of content most consumed.

## Materials and Methods

A cross-sectional study was conducted between January 2020 – February 2021 [barring the period from March 2020 to October 2020 – due to the COVID-19 lockdown], in the districts of Mumbai City, Mumbai Suburban, Thane and Palghar, wherein urban and rural units were considered. Sample size 400 was calculated using Cochran’s formula; n ≥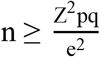assuming maximum variability, i.e., equal to 50% (p = 0.5), 95% confidence level with ± 5% precision considered.

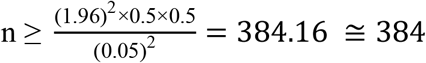, rounding off to 400.

The Sample was stratified in appropriate proportions based on age groups according to the Census of India 2011 to maintain population representativeness as depicted in Table 1.

**Table 1:**
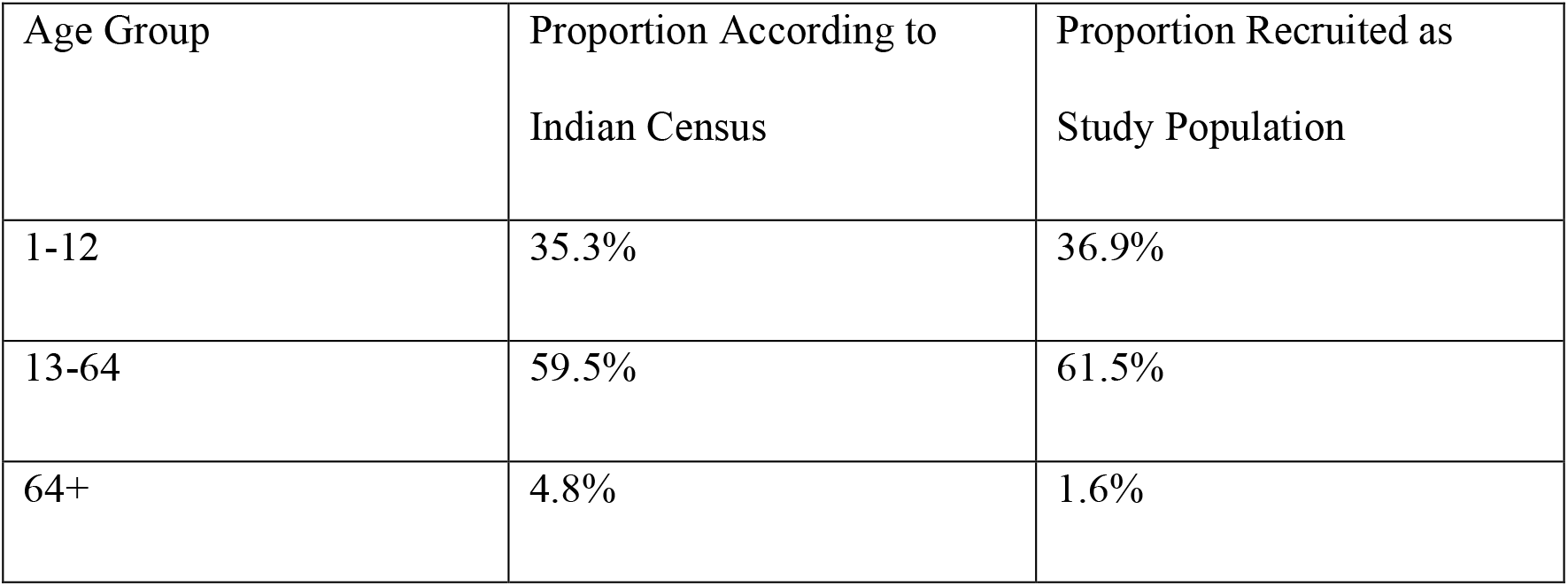
Proportion of participants recruited based on age group.

Analysis of data of age group 1-12 was done separately and only the results of population above the age of 13 (252 participants) have been presented in this article. Stratified random sampling was used wherein, individuals of the age group 13-24 years were recruited from 3 schools and 5 colleges by selecting random roll numbers, whereas individuals of the age groups 25 and above were recruited from 2 urban & 2 rural housing societies by selecting random house numbers. These numbers were generated using a random number generating website. Individuals who had access to electronic screen devices & were capable of comprehension and verbal communication were included, whereas those with terminal illness, critical conditions or a history of severe psychiatric disorder were excluded. The questionnaire was administered in a language that the respondents could best comprehend and data was recorded simultaneously.

### 3.1 Ethical considerations

The study commenced after being approved by the Institutional Ethics Committee (EC/OA-133/2019). All the procedures followed were in accordance with the ethical standards of the responsible committee on human experimentation (institutional or regional) and with the Helsinki Declaration of 1975, as revised in 2000. Informed consent was obtained from individuals above 18 years of age and written assent from individuals between age 13-17 years along with informed consent from their parents.

### 3.2 Study Tools

#### 1. Digital Addiction Scale

The DAS, comprising of 19 items, each with a five-point Likert scale, was administered to measure addiction level of the participants. It has a Cronbach’s alpha reliability of the of 0.874.^[17]^ Criterion-based validity of the DAS has been determined by applying the DAS concurrently with the YIATSF, SPAS-SF, DGAS, and FAS, gave correlation coefficients as 0.833, 0.756, 0.600, and 0.447 (all of them are significant at the 0.001 level) successively ^[17]^. Final DAS score is calculated by dividing the total score by 19. Thus, the scoring range of the scale is from 1.00 to 5.00, where higher score indicated greater screen dependency. Due to the lack of cut-offs provided by copyright owner of the scale, a pilot study among 30 participants was conducted after which the following cut-offs were considered as interpretation criteria, with the consultation with a panel of 5 psychiatrists. The cut off were, “1.00-2.49”-Normal, “2.50-3.49”-Problematic Screen User & “3.50-5.00”-Addicted Screen User.

#### 2. Questionnaire to assess Screen Usage Behaviour & Content Consumed

Questionnaire included multiple choice questions & Yes/No type questions to assess the amount of screen time, screen use behaviours, types of content consumed, withdrawal symptoms and screen use specific psychological consequences. The Questionnaire was validated by the 5 experts from the Department of Psychiatry. The content validity ratio of the Questionnaire was > 0.65. Both the study tools were translated and made available in local languages (Hindi & Marathi).

#### 3.3 Statistical Analysis

The data was entered in an Excel sheet and analysed using the IBM Corp. Released 2019. IBM SPSS Statistics for Windows, Version 26.0. Armonk, NY: IBM Corp. The calculations were carried out in numbers and their percentages. Chi-square test was used for analysing the association of screen addiction with categorical variables. Due to the ordinal nature of the DAS Score, Kruskal Wallis test was used to compare the addiction scores with respect to type of device used & content consumed. Paired t test was used to compare routine and holiday screen time. All *P* values <0.05 were considered statistically significant.

## 4. Results

### 4.1 Demographics

Responses were obtained from a total of 252 participants out of which 54% were males and 46% were females. 44.4 % belonged to the age group of 13-25, 35.3 % to 26-44, 17.9% to 45-65 & 2.4% above 65 years of age. 5.9% participants had a level of education up to the matric level, 21.8% up to high school level, 19.8% were graduates and 41.3% were postgraduates. 75% participants were residents of urban areas while the rest 25% were from the rural areas. Approximately equal representation was obtained from Schools & Colleges (Degree, Engineering & Medical). Maximum responses were obtained from students and service employees (39.7% and 29% respectively).

### 4.2 Screen Time

Routine Screen time (excluding profession and educational use) was found to be 2.99 ±2.01 hrs which increased significantly during the holidays to 4.45 ±2.02 hrs (p=0.001). Mean longest during of continuous screen was found to be 3.60 ± 2.75 hrs, the upper limit of which was as high as 18hrs. Median screen free duration found to be just 8hrs.

### 4.3 Prevalence of Screen Addiction

The Prevalence of Screen addiction was found to be 13.1% in the general population. Problematic users were found to be 34.5% and the rest 52% users were normal. Maximum prevalence of screen addiction was found in the age group of 13-25 (23.2%) and males (p=0.001). There was no significant difference in the prevalence with respect to the region of residence. Table 2.

**Table 2:** Association of Screen Addiction with Demographic Variables.

| Variable |  | Normal<br>% (n) | Problematic<br>% (n) | Addict<br>% (n) | Total<br>(N=252) | Pearson<br>chi<br>square<br>value | df | P value |
| --- | --- | --- | --- | --- | --- | --- | --- | --- |
| Age group | 13-25 | 38.4% (43) | 38.4% (43) | 23.2% (26) | 112 | 28.296 | 6 | 0.001* |
|  | 26-44 | 59.6% (53) | 34.8% (31) | 5.6% (5) | 89 |  |  |  |
|  | 45+ | 70.6% (36) | 25.5% (13) | 3.9% (2) | 51 |  |  |  |
| Sex | Female | 64.7% (75) | 24.1% (28) | 11.2% (13) | 116 | 13.483 | 2 | 0.001* |
|  | Male | 41.9% (57) | 43.4% (59) | 14.7% (20) | 136 |  |  |  |
|  | Rural | 42.9% (27) | 41.3% (26) | 15.9% (10) | 63 | 3.057 | 2 | 0.217 |

| Region of residence | Urban | 55.6% (105) | 32.3% (61) | 12.2% (23) | 189 |  |  |  |
| --- | --- | --- | --- | --- | --- | --- | --- | --- |
| School/College | Medical College | 5.2% (1) | 36.8% (7) | 57.9% (11) | 19 | 27.87 | 6 | 0.001* |
|  | Engineering College | 45.5% (10) | 40.9% (9) | 13.6% (3) | 22 |  |  |  |
|  | Other College | 29.6% (8) | 55.5% (15) | 14.8% (4) | 27 |  |  |  |
|  | School | 60.7% (17) | 107% (3) | 28.6% (8) | 28 |  |  |  |
| * p<0.05, statically significant. Percentages are taken out of the total of each row |  |  |  |  |  |  |  |  |

The addicts were further classified into social media addicts (9.1%) binge watching addicts (2.3%), and gaming addicts (1.2%) based on their most consumed content. Figure 1. The difference in their prevalence in was statistically significant (p=0.001). The prevalence of screen addiction and problematic screen usage was also significantly higher in those who used smartphones the most (p= 0.016) compared to other devices. Table 3.

**Figure 1:**
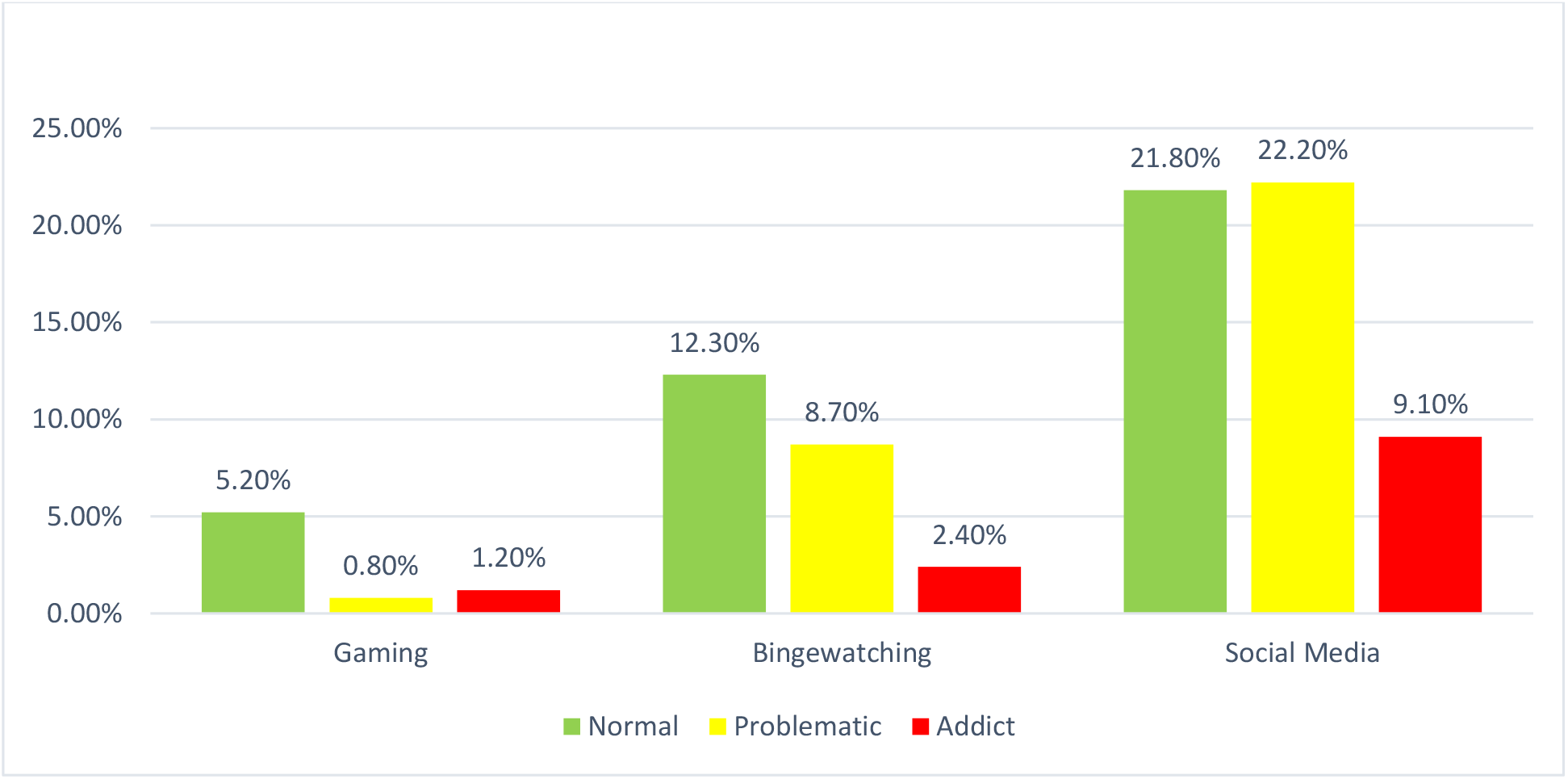
Prevalence of screen addiction based on the most consumed content.

**Table 3:** Association of Screen Addiction with Most used Device & Most Consumed Content.

| Variable |  | Normal<br>% (n) | Problematic<br>% (n) | Addict<br>% (n) | Total<br>(N=252) | Median<br>DAS<br>score | P<br>value |
| --- | --- | --- | --- | --- | --- | --- | --- |
| **Most used**<br>**Device** | Laptop | 76.5% (13) | 17.6% (3) | 5.9% (1) | 17 | 1.89 | 0.001* |
|  | PC | 71.4% (10) | 21.4% (3) | 7.1% (1) | 14 | 1.89 |  |
|  | Smartphone | 46.0% (91) | 38.9% (77) | 15.2% (30) | 198 | 2.63 |  |
|  | TV | 78.3% (18) | 17.4% (4) | 4.3% (1) | 23 | 1.73 |  |
| **Most**<br>**Consumed**<br>**Content** | Binge watching (TV series/<br>Movies/ Entertainment videos) | 52.5% (31) | 37.7% (22) | 10.2% (6) | 59 | 2.42 | 0.001* |
|  | Gaming | 72.2% (13) | 11.1% (2) | 16.7% (3) | 18 | 2.21 |  |
|  | Social Media | 41.0% (55) | 41.8% (56) | 17.2% (23) | 134 | 2.68 |  |
|  | Other | 80.5% (33) | 17.1% (7) | 2.4% (1) | 41 | 1.89 |  |

\* p<0.05, statically significant
Percentages are taken out of the total of each row

The addiction scores of participants with respect to most used device and most consumed was found to be as depicted in Table 3. Median score of those who used smartphones the most was found to be significantly higher as compare to those who preferred to used other devices(p=0.001). With regard to the most consumed content the median scores of social media users, gamers and binge-watchers were significantly higher than those who preferred to use screen devices for any other purpose(p=0.001). However, when one to one comparison of these scores was performed, no significant difference between gaming-binge watching, gaming-social media, binge watching -social media was found indicating that degree of addiction is similar for all the three types of content(p>0.05).

### 4.4 Association with self-reported motivation of frequent screen use

The association of screen addiction with the self-reported motivations of screen use is as depicted in the Table 4. The prevalence of screen addiction and problematic screen usage was significantly higher in those who frequently used screens devices for motivations like, watching relatable content (p= 0.005), watching violent content (p= 0.006), watching nudity (p= 0.003), peer pressure (p= 0.001), popularity of game/tv show/app (p=0.001), to socialise with new people over a common topic(p=0.009), availability of ample free time (p=0.014), get over loneliness (p=0.001), get over depression (p=0.001), get over stress (p=0.001). The proportion of users who used screen devices more frequently for educational content (p=0.001) was significantly higher in normal group.

**Table 4.**
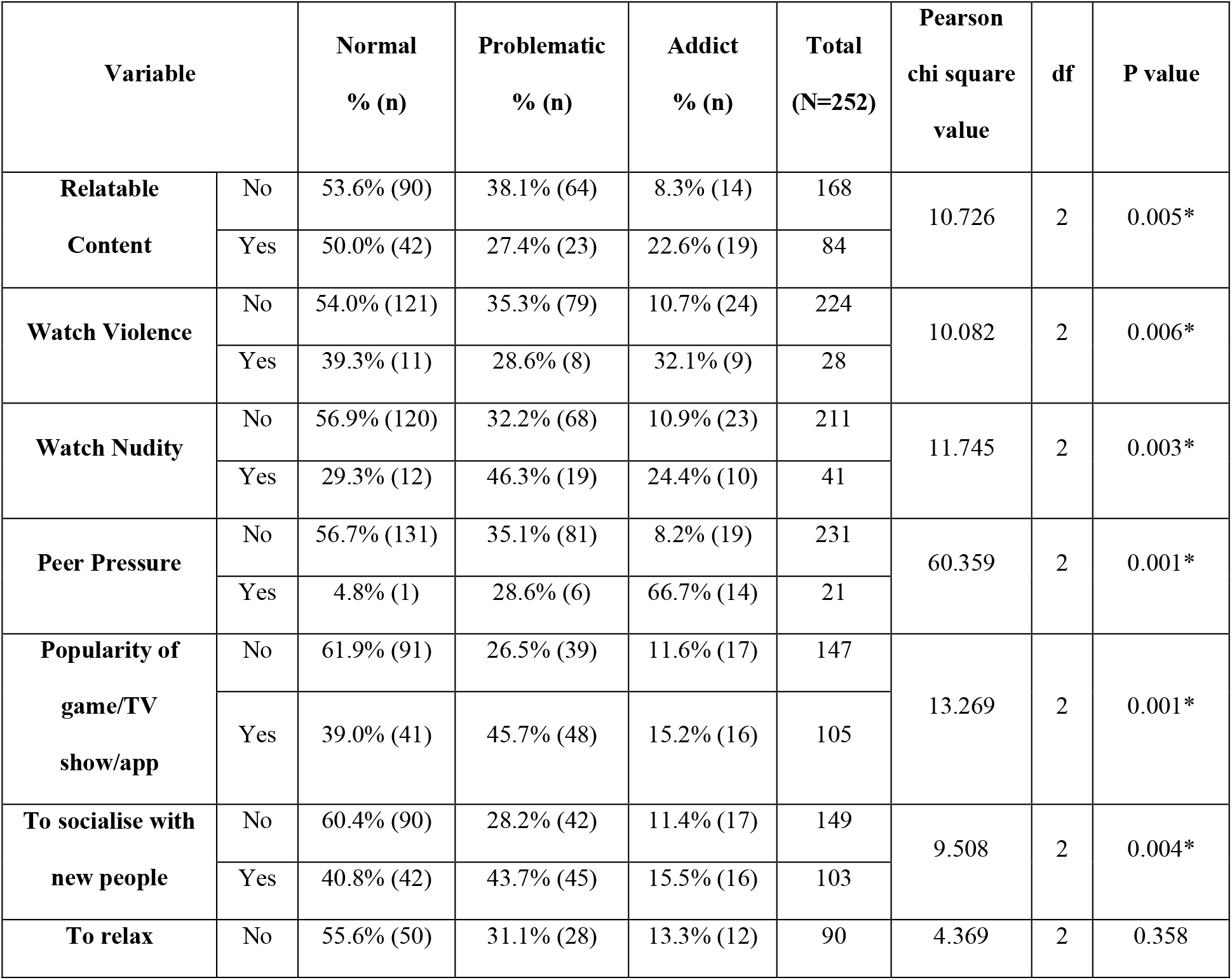
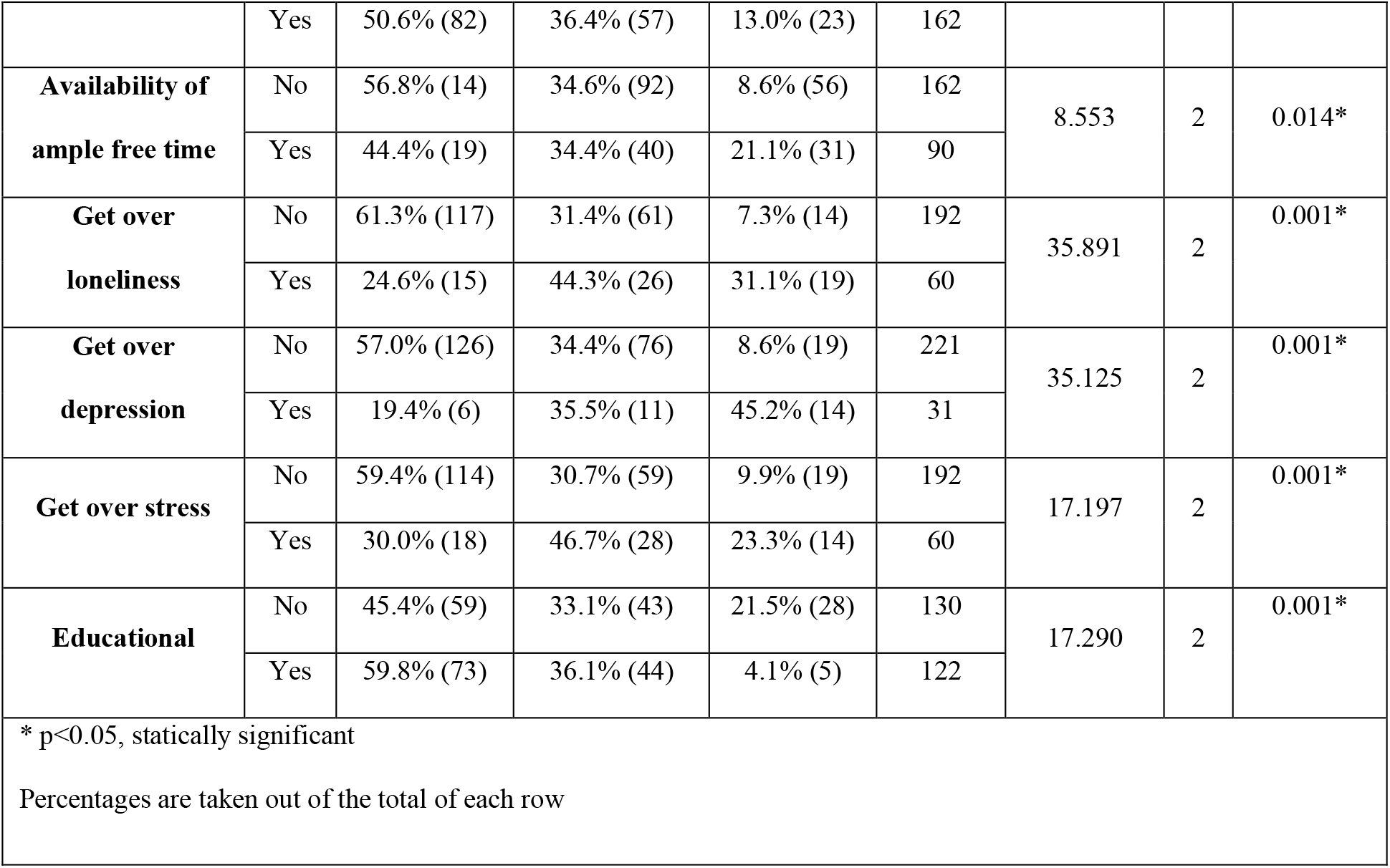
Association of Screen Addiction with self-reported Motivations of frequent Screen use.

| Variable |  | Normal<br>% (n) | Problematic<br>% (n) | Addict<br>% (n) | Total<br>(N=252) | Pearson<br>chi square<br>value | df | P value |
| --- | --- | --- | --- | --- | --- | --- | --- | --- |
| Relatable<br>Content | No | 53.6% (90) | 38.1% (64) | 8.3% (14) | 168 | 10.726 | 2 | 0.005* |
|  | Yes | 50.0% (42) | 27.4% (23) | 22.6% (19) | 84 |  |  |  |
| Watch Violence | No | 54.0% (121) | 35.3% (79) | 10.7% (24) | 224 | 10.082 | 2 | 0.006* |
|  | Yes | 39.3% (11) | 28.6% (8) | 32.1% (9) | 28 |  |  |  |
| Watch Nudity | No | 56.9% (120) | 32.2% (68) | 10.9% (23) | 211 | 11.745 | 2 | 0.003* |
|  | Yes | 29.3% (12) | 46.3% (19) | 24.4% (10) | 41 |  |  |  |
| Peer Pressure | No | 56.7% (131) | 35.1% (81) | 8.2% (19) | 231 | 60.359 | 2 | 0.001* |
|  | Yes | 4.8% (1) | 28.6% (6) | 66.7% (14) | 21 |  |  |  |
| Popularity of<br>game/TV<br>show/app | No | 61.9% (91) | 26.5% (39) | 11.6% (17) | 147 | 13.269 | 2 | 0.001* |
|  | Yes | 39.0% (41) | 45.7% (48) | 15.2% (16) | 105 |  |  |  |
| To socialise with<br>new people | No | 60.4% (90) | 28.2% (42) | 11.4% (17) | 149 | 9.508 | 2 | 0.004* |
|  | Yes | 40.8% (42) | 43.7% (45) | 15.5% (16) | 103 |  |  |  |
| To relax | No | 55.6% (50) | 31.1% (28) | 13.3% (12) | 90 | 4.369 | 2 | 0.358 |
|  | Yes | 50.6% (82) | 36.4% (57) | 13.0% (23) | 162 |  |  |  |
| Availability of ample free time | No | 56.8% (14) | 34.6% (92) | 8.6% (56) | 162 | 8.553 | 2 | 0.014* |
|  | Yes | 44.4% (19) | 34.4% (40) | 21.1% (31) | 90 |  |  |  |
| Get over loneliness | No | 61.3% (117) | 31.4% (61) | 7.3% (14) | 192 | 35.891 | 2 | 0.001* |
|  | Yes | 24.6% (15) | 44.3% (26) | 31.1% (19) | 60 |  |  |  |
| Get over depression | No | 57.0% (126) | 34.4% (76) | 8.6% (19) | 221 | 35.125 | 2 | 0.001* |
|  | Yes | 19.4% (6) | 35.5% (11) | 45.2% (14) | 31 |  |  |  |
| Get over stress | No | 59.4% (114) | 30.7% (59) | 9.9% (19) | 192 | 17.197 | 2 | 0.001* |
|  | Yes | 30.0% (18) | 46.7% (28) | 23.3% (14) | 60 |  |  |  |
| Educational | No | 45.4% (59) | 33.1% (43) | 21.5% (28) | 130 | 17.290 | 2 | 0.001* |
|  | Yes | 59.8% (73) | 36.1% (44) | 4.1% (5) | 122 |  |  |  |
| * p<0.05, statically significant |  |  |  |  |  |  |  |  |
| Percentages are taken out of the total of each row |  |  |  |  |  |  |  |  |

### 4.5 Association with types of apps used and genres played/watched

The association of screen addiction with the type of application used & genre played/watched is as depicted in Table 5. The prevalence of screen addiction and problematic screen usage was significantly higher among those who used Instagram (p=0.030), Snapchat (p= 0.001), played shooter games (p= 0.042), sports games (p=0.038), used Amazon Prime Video(p=0.015), YouTube (p= 0.001), used Offline resources to watch TV Shows/Movies (p= 0.001) and watched the genres of Crime -Suspense(p=0.004) & action-adventure (p=0.002) TV Shows/Movies.

**Table 5:**
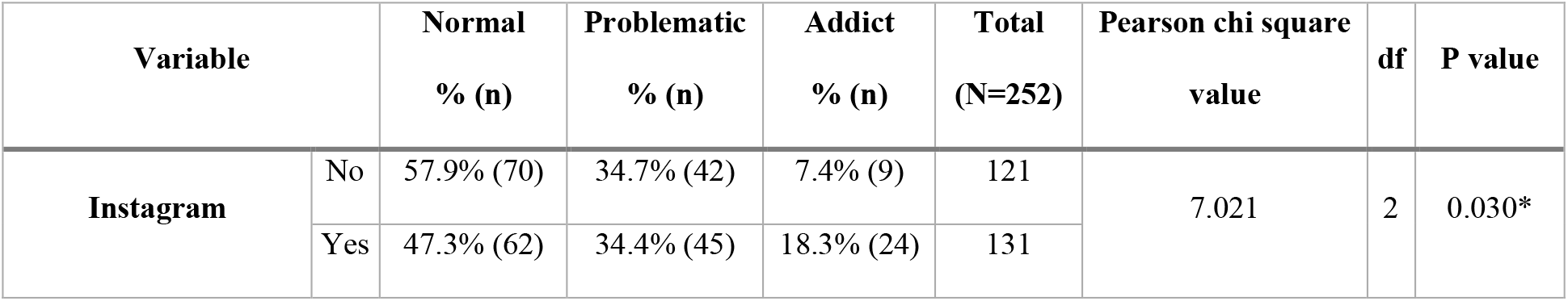
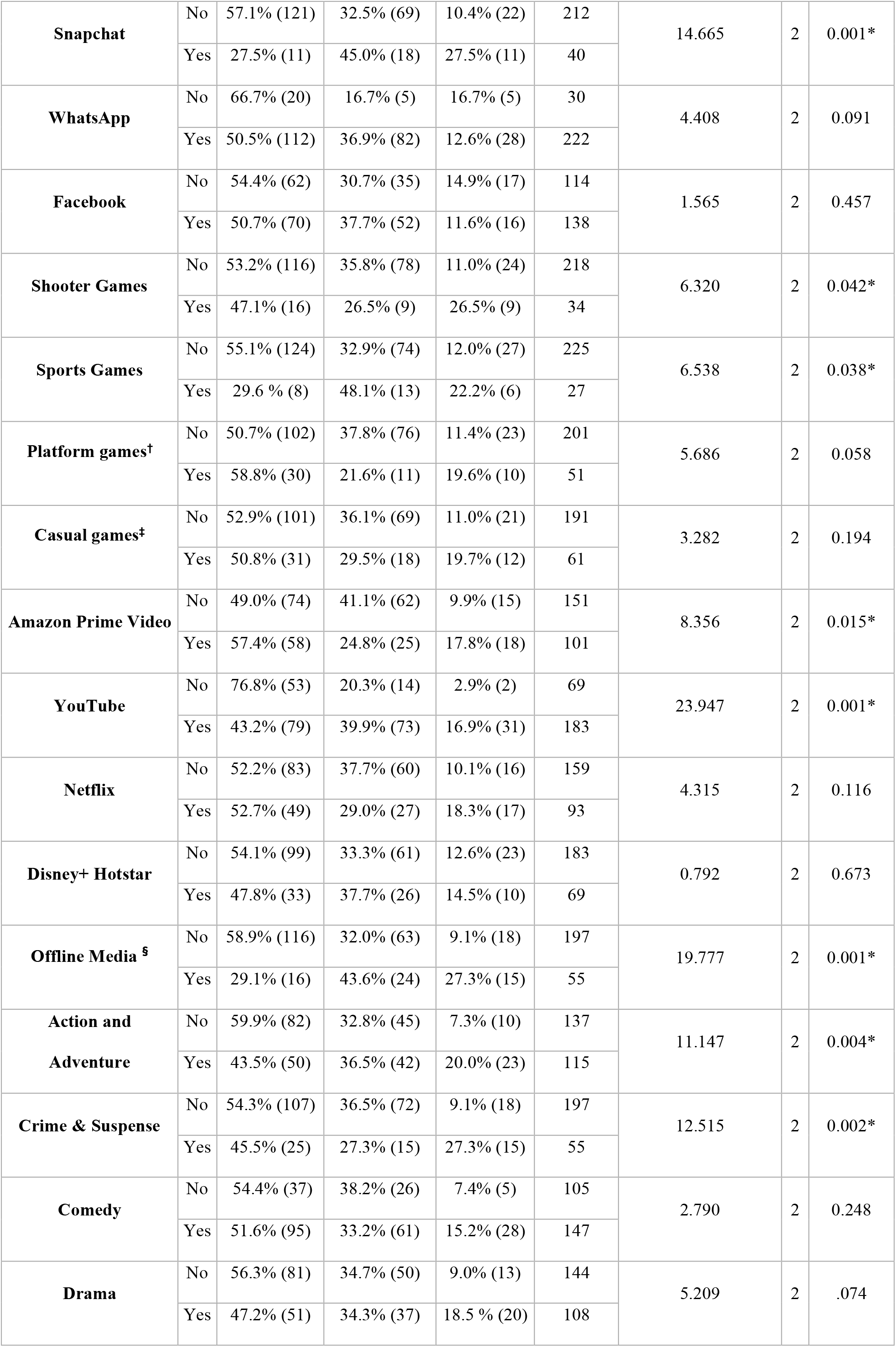
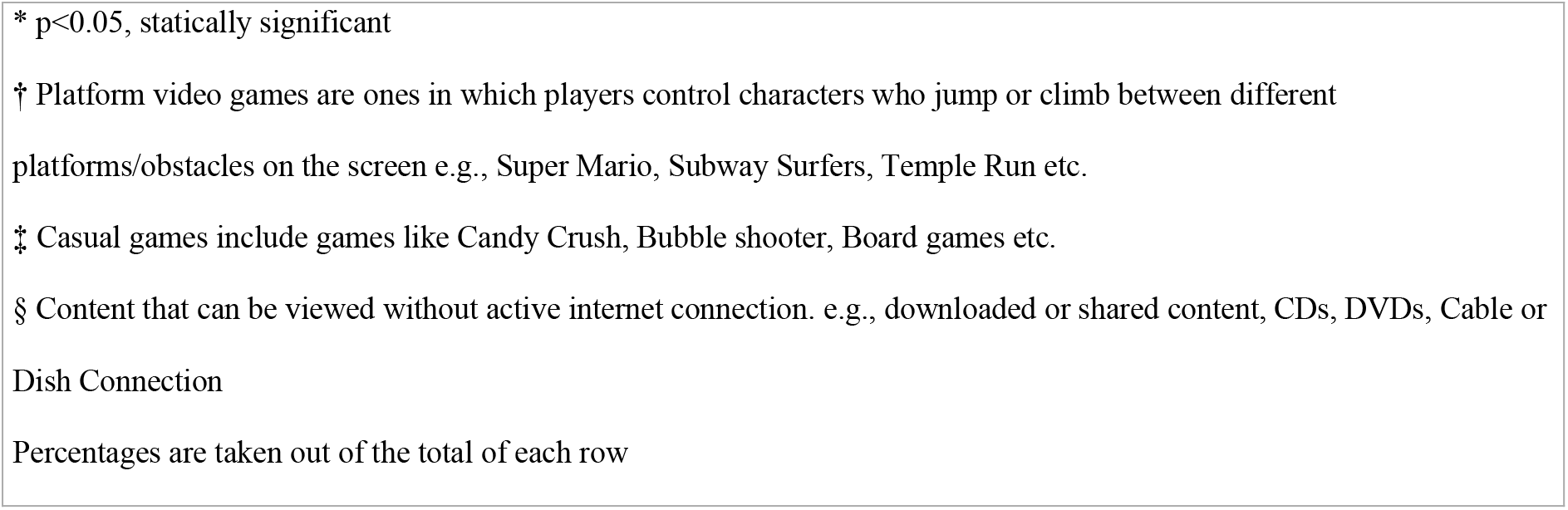
Association of screen addiction with types of apps used and genres played/watched.

| Variable |  | Normal<br>% (n) | Problematic<br>% (n) | Addict<br>% (n) | Total<br>(N=252) | Pearson chi square<br>value | df | P value |
| --- | --- | --- | --- | --- | --- | --- | --- | --- |
| **Instagram** | No | 57.9% (70) | 34.7% (42) | 7.4% (9) | 121 | 7.021 | 2 | 0.030* |
|  | Yes | 47.3% (62) | 34.4% (45) | 18.3% (24) | 131 |  |  |  |
| **Snapchat** | No | 57.1% (121) | 32.5% (69) | 10.4% (22) | 212 | 14.665 | 2 | 0.001* |
|  | Yes | 27.5% (11) | 45.0% (18) | 27.5% (11) | 40 |  |  |  |
| **WhatsApp** | No | 66.7% (20) | 16.7% (5) | 16.7% (5) | 30 | 4.408 | 2 | 0.091 |
|  | Yes | 50.5% (112) | 36.9% (82) | 12.6% (28) | 222 |  |  |  |
| **Facebook** | No | 54.4% (62) | 30.7% (35) | 14.9% (17) | 114 | 1.565 | 2 | 0.457 |
|  | Yes | 50.7% (70) | 37.7% (52) | 11.6% (16) | 138 |  |  |  |
| **Shooter Games** | No | 53.2% (116) | 35.8% (78) | 11.0% (24) | 218 | 6.320 | 2 | 0.042* |
|  | Yes | 47.1% (16) | 26.5% (9) | 26.5% (9) | 34 |  |  |  |
| **Sports Games** | No | 55.1% (124) | 32.9% (74) | 12.0% (27) | 225 | 6.538 | 2 | 0.038* |
|  | Yes | 29.6 % (8) | 48.1% (13) | 22.2% (6) | 27 |  |  |  |
| **Platform games<sup>†</sup>** | No | 50.7% (102) | 37.8% (76) | 11.4% (23) | 201 | 5.686 | 2 | 0.058 |
|  | Yes | 58.8% (30) | 21.6% (11) | 19.6% (10) | 51 |  |  |  |
| **Casual games<sup>‡</sup>** | No | 52.9% (101) | 36.1% (69) | 11.0% (21) | 191 | 3.282 | 2 | 0.194 |
|  | Yes | 50.8% (31) | 29.5% (18) | 19.7% (12) | 61 |  |  |  |
| **Amazon Prime Video** | No | 49.0% (74) | 41.1% (62) | 9.9% (15) | 151 | 8.356 | 2 | 0.015* |
|  | Yes | 57.4% (58) | 24.8% (25) | 17.8% (18) | 101 |  |  |  |
| **YouTube** | No | 76.8% (53) | 20.3% (14) | 2.9% (2) | 69 | 23.947 | 2 | 0.001* |
|  | Yes | 43.2% (79) | 39.9% (73) | 16.9% (31) | 183 |  |  |  |
| **Netflix** | No | 52.2% (83) | 37.7% (60) | 10.1% (16) | 159 | 4.315 | 2 | 0.116 |
|  | Yes | 52.7% (49) | 29.0% (27) | 18.3% (17) | 93 |  |  |  |
| **Disney+ Hotstar** | No | 54.1% (99) | 33.3% (61) | 12.6% (23) | 183 | 0.792 | 2 | 0.673 |
|  | Yes | 47.8% (33) | 37.7% (26) | 14.5% (10) | 69 |  |  |  |
| **Offline Media <sup>§</sup>** | No | 58.9% (116) | 32.0% (63) | 9.1% (18) | 197 | 19.777 | 2 | 0.001* |
|  | Yes | 29.1% (16) | 43.6% (24) | 27.3% (15) | 55 |  |  |  |
| **Action and Adventure** | No | 59.9% (82) | 32.8% (45) | 7.3% (10) | 137 | 11.147 | 2 | 0.004* |
|  | Yes | 43.5% (50) | 36.5% (42) | 20.0% (23) | 115 |  |  |  |
| **Crime & Suspense** | No | 54.3% (107) | 36.5% (72) | 9.1% (18) | 197 | 12.515 | 2 | 0.002* |
|  | Yes | 45.5% (25) | 27.3% (15) | 27.3% (15) | 55 |  |  |  |
| **Comedy** | No | 54.4% (37) | 38.2% (26) | 7.4% (5) | 105 | 2.790 | 2 | 0.248 |
|  | Yes | 51.6% (95) | 33.2% (61) | 15.2% (28) | 147 |  |  |  |
| **Drama** | No | 56.3% (81) | 34.7% (50) | 9.0% (13) | 144 | 5.209 | 2 | .074 |
|  | Yes | 47.2% (51) | 34.3% (37) | 18.5 % (20) | 108 |  |  |  |

\* $p < 0.05$ , statically significant
† Platform video games are ones in which players control characters who jump or climb between different platforms/obstacles on the screen e.g., Super Mario, Subway Surfers, Temple Run etc.
‡ Casual games include games like Candy Crush, Bubble shooter, Board games etc.
§ Content that can be viewed without active internet connection. e.g., downloaded or shared content, CDs, DVDs, Cable or Dish Connection
Percentages are taken out of the total of each row

### 4.6 Association with withdrawal symptoms

One of the major criteria to define any behaviour as addictive is presence of withdrawal symptoms. The prevalence of withdrawal symptoms like anxiety (p=0.002), desperation to use the device again (p= 0.001), depression (p=0.007), irritation and aggression (p= 0.001), and loss of concentration (p=0.009) was significantly higher among addicts and problematic screen users.

### 4.7 Association with Screen use specific psychological consequences

Our study also reported screen use specific psychological consequences like:

- Texting (26.2%) & Ringing (15.47%) Anxiety – urge to repeatedly check the phone & sensing phantom vibrations ^[18]^
- Selfitis (18.65%) -urge to click multiples selfies to cope up with low self-esteem & body image ^[19]^
- Para-Social Relationships (12.3%)-forming emotional relationships with virtual/fictional characters ^[20]^
- Game Transfer Phenomenon (17.3%) -temporarily seeing images, hearing music, sounds, voices, tactile sensations, involuntary movements of limbs, illogical thoughts, verbal outbursts, even when not playing the video game ^[21]^
- Episode Transfer Phenomenon (8.73%)-symptoms similar to game transfer but in context of TV series which has been reported for the first time among the binge watcher through our study.

All these psychological consequences were found to be significantly associated with screen addiction (p= 0.001).

## 5. Discussion

The objective of our study was to determine the prevalence of screen addiction and its association with screen use behaviour and type of content consumed. The prevalence of screen addiction was found to be 13.1% and was significantly higher amongst 13-25 age group (school and college-based population). Epidemiological studies have reported a significant variation in the prevalence rates among adolescents and young people from 6.3 to 37.9% in Asia.^[22]^ Among general population of India, the prevalence was found to be 1.3%.^[11]^ Higher rates of 11.8%, 8.8%, and 8% have been reported in college populations. ^[12, 13, 14]^ Our results are consistent with previous Asian studies in which the prevalence has been found higher in males & college-based population.^[23]^ However, we found no significant difference amongst urban & rural populations whereas the previous studies did find a significantly higher prevalence in urban areas.^[22]^ Ours is one of the very few studies which has taken older adults into consideration & has estimated the prevalence by recruiting proportionate number of participants according to general population demographics. The major reason for the disparities in prevalence remains lack of an agreed upon criteria to define & diagnose Internet addiction. Various classifications of Internet addiction have been proposed. For instance, Young and colleagues grouped five different forms of addictive behaviour i.e., the use of computer itself, information searching, cyber sexuality, cyber contracts, and net compulsions like gaming and shopping addictions, etc. ^[24]^ Other terms which have been used in the context of screen-based addiction are Smartphone Addiction ^[25]^ & TV addiction ^[26]^ which have tried to address the offline component of problematic & addictive Screen use. However, it is only gaming disorder which has been recognized by WHO in the ICD-11. ^[27]^

We attempted to classify screen addiction into Gaming, social media & Binge-watching using a scale which took both online & offline usage into consideration & items of which were based on overall screen use. However, we didn’t take into account cyber sexuality & online shopping. We found that there was no significant difference between the median DAS (Digital Addiction Scale) scores of social media users, gamers and binge-watchers. Considering the most used device, our study found scores of mobile users to be significantly high while previous studies have found association of addictive behaviours with laptops, PCs & TVs through which content like games, movies & TV shows can be accessed. ^[26, 28]^

Evidence suggests that excessive binge watching is similar to other related addictive behaviours like video gaming, internet or social media addiction. People binge-watch to connect socially, to become a part of any fandom group, and under peer-pressure. Studies show that increased ‘Fear of missing out (FOMO)’ was a significant predictor for the phenomenon of binge-watching. The other psychological motivations resulting in binge-watching are the urge to escape from daily life problems, coping with loneliness, stress & depression. ^[29]^ In our study, all of these factors were found to be significantly associated with problematic & addictive screen use not only amongst binge watchers but also amongst gamers & social media users. Previous studies have also found association of such kind of screen use behaviour with social media ^[30]^ & gaming addiction ^[28]^ but studies associating such behaviour with addictive binge-watching are very few. One of the novel findings of our study was the association of screen addiction with motivation to watch violent content & nudity which is increasingly becoming a part of many shows & movies available on the OTT platforms.

Regarding the applications used by the participants, we found Instagram & Snapchat to be to associated with addiction, but not Facebook, WhatsApp & Twitter. Previous studies have found Facebook to be associated with addiction ^[31]^ but considering the change in trends, today Facebook is a platform for older adults whereas Instagram & Snapchat have become more popular among the youth whose tendency to get addicted is significantly higher. ^[30]^ Amongst the OTT & Streaming platforms only YouTube & Amazon Prime Video were found to be associated with addiction, however there have been reports suggesting other platforms like Netflix, Disney+ Hotstar, etc also design their algorithms & promotions such that they favour the practice of Binge-watching. ^[32, 33]^ The strong association of addiction with watching content by either downloading or sharing offline, which is practiced by users not having subscription to OTT platforms also highlights the importance of taking offline screen use into account while defining screen addiction. The association of addiction with genres of games like Shooting & that of TV Shows/Movies like Crime-Suspense & action-adventure is a matter of concern as these genres depict violence content. High exposure to violent content has proven to influence the behaviour of the individual, making them more impatient, aggressive & violent which also frequently manifests as a withdrawal behaviour. ^[34]^ While Withdrawal symptoms have been previously reported in gaming ^[35]^ & social media addicts, ^[30]^ reporting withdrawal amongst the Binge-watchers is again one of the novel findings of our study.

Our study also found association of screen addiction with screen use specific psychological consequences like, Texting & Ringing Anxiety ^[18]^ – urge to repeatedly check the phone & sensing fantom vibrations; Selfitis ^[19]^-urge to click multiples selfies to cope up with low self-esteem & body image; Para-Social Relationships-forming emotional relationships with virtual/fictional characters. ^[20]^; Game Transfer Phenomenon ^[21]^-seeing images, hearing sounds and feeling tactile unreal sensations temporarily, involuntary movement of limbs, verbal outbursts even when not playing the video game; Episode Transfer Phenomenon-symptoms similar to game transfer but in context of TV series which has been reported for the first time amongst the binge watcher through our study.

King et al. in their study mention that during the treatment of gaming addiction when the users were abstained from playing certain video games, they switched to binge-watching the gameplay (videos of those games) on YouTube, which highlights the possibility of an addict switching the mode through which content is being consumed & also of co-existence of addiction to multiple types of content.^[36]^ With the changing trend on how screen devices are being used, there is need to take into consideration that a user can use multiple devices to watch the same content as well as use a single device to consume multiple types of content, both online & offline. Hence, there is a need to revise the exiting classification which focuses only on internet use & bring all kinds of addictive screen usage under a broader umbrella of ‘Screen Addiction’. A standardised diagnostic criterion could be made on similar lines by conducting more longitudinal clinical studies. Individuals also need to be made aware about moderating the consumption of addictive & violent content as well as also avoid relying on screens for the self-reported motivations which are found to be addictive.

## Limitations

Since data from housing societies was collected after the 1^st^ COVID-19 lockdown, which saw a significant rise in the screen use, there is a likelihood that some of our results might be skewed. Due to the cross-sectional nature of the study, causation of screen addiction with associated risk factors could not be established. Authors have not considered cyber sexuality, compulsive internet gambling and shopping under screen addiction. The sample though representative, is relatively small and future studies involving multiple cities and villages can be undertaken.

## 6. Conclusion

The prevalence of screen addiction is substantially high, particularly in the younger population. This study also highlights the strong association of screen use behaviour, type of content consumed and apps used with addictive screen use. The similarity in addiction scores and associated factors also support the use of umbrella term ‘Screen Addiction’ to group all kinds of addictive screen usage.

## Data Availability

All data produced in the present study are available upon reasonable request to the authors

